# Scoping into Non Iron Deficiency Anemia: Reflection from the Rural India

**DOI:** 10.1101/2021.02.10.21251472

**Authors:** Somen Saha, Tapasvi Puwar, Deepak Saxena, Komal Shah, Apurva kumar Pandya, Nikita Vadsariya, Bharat Desai, Raj Suthariya

## Abstract

**Introduction:** Anaemia is one of the leading public health problems. India accounts for the highest prevalence of anaemia in the world. Anaemia programs in India focus on screening and management of anaemia based on haemoglobin estimation, treatment is being given irrespective of status of iron as well as other micronutrient storage. The present study assesses the prevalence of anaemia and iron deficiency (ID) based on low serum ferritin status among antenatal and postnatal women in Devbhoomi Dwarka District of Gujarat.

**Methods:** A total of 258 pregnant (AN) and postnatal (PN) women drawn from 27 primary health centres were studied. Anaemia was evaluated based on haemoglobin concentration obtained from venous whole blood, using auto-analyser. Serum ferritin was used to evaluate iron status in the study. Serum ferritin was assessed using the direct chemiluminescence method using MINI VIDAS which is a compact automated immunoassay system based on the Enzyme Linked Fluorescent Assay (ELFA) principles.

**Results:** Overall, Anaemia (low Hb) and ID (low s. ferritin) was observed in 65.9% and 27.1% respectively. Out of anaemic participants, about 38.2% reported ID while the remaining 61.8% had normal s. ferritin (i.e. non-iron deficient anaemia). Anaemia was reported 69.1% in AN women and 57.1% in PN women. The ID was reported higher (30.9%) in AN woman than PN women (17.1%). However, the prevalence of anaemia, as well as IDA decreased from the first to the third trimester.

**Conclusion:** Two out of every three women were anaemic; one out of four were anaemic with depleted iron storage. Importantly, two out of five women had anaemia but iron storage was sufficient. Strategy to prevent and correct anaemia must include screening for iron and non-iron deficiency anaemia and follow appropriate treatment protocol for both types of anaemia.

## Introduction

Anaemia is one of the common public health problems in India. According to The National Family Health Survey (2016) ^[1]^, 58.6% of children, 53.2% of non-pregnant women and 50.4% of pregnant women were anaemic.

Iron deficiencies are the most common cause of anaemia in the world. ^[2]^ Other micronutrients such as folate, B12 and vitamin A and infections such malaria, TB, worm infestation, haemorrhoids and other inherited diseases of haemoglobin synthesis are also causes of Anaemia, which may complicate the ante and post-natal period. ^[3]^

Presently in India, under the Intensive National Iron Plus Initiative (INIPI), ^[4]^ oral and parenteral iron preparation, folate and albendazole are emphasized for prevention and treatment of maternal anaemia. Also, food fortification and food diversity are promoted as a strategy to prevent nutritional anaemia. Despite these efforts, anaemia is still a problem of public health.

Although Iron Deficiency (ID) is the reason for anaemia in 50% women worldwide but among half of the anaemic condition is other than ID. ^[2]^ However, as per current protocol, screening and management of maternal anaemia is planned by Haemoglobin (Hb) estimation^[4]^ only, and treatment is being given irrespective of the status of iron as well as other micronutrient storage. Existing literature on IDA and Non-Iron Deficiency Anaemia (NIDA) are in the global context; however, studies on this topic is limited in Indian context. The present study assesses the prevalence of anaemia and iron deficiency (ID) based on low serum ferritin status among antenatal and postnatal women in Devbhoomi Dwarka District of Gujarat.

## Methods

A total of 258 pregnant (AN) and postnatal (PN) women drawn from 27 primary health centres were studied. Anaemia was assessed based on haemoglobin concentration obtained from venous whole blood, using auto-analyser. Serum ferritin was used to assess iron status in the study. Serum ferritin was assessed using the direct chemiluminescence method using MINI VIDAS which is a compact automated immunoassay system based on the Enzyme Linked Fluorescent Assay (ELFA) principles.

The pregnant women of any trimester (Antenatal – AN Women) and postnatal women (PN Women) within 90 days of pregnancy were included in the present study. After taking informed consent, basic details of participants such as name, age, gender, residence and obstetric history were gathered through pre-tested semi-structured questionnaires. From every participant 4 ml venous blood samples were taken as per the standard procedure through phlebotomist, lab technicians, at the respective PHC. The specimens were transported to the laboratory within one hour of collection and were stored at −20°C until the time of analysis.

Anaemia was defined as hemoglobin <11% in AN women and <12% in PN women. ^[5]^ IDA is defined as a combination of anemia and ID while NIDA is defined as the presence of anemia without ID. Low serum ferritin with normal C-reactive protein level was considered as a biomarker for iron deficiency. S. ferritin as a more sensitive and specific marker for ID estimation and for planning management of maternal anemia.^[6,7]^ Similar marker was used by Comprehensive National Nutritional Survey-2018-19 ^[8]^, India. Following the WHO guidelines, ID based on s. ferritin level was defined as <12 ng/ml in AN women and <15ng/dl in PN Women ^[9,10]^ S. ferritin levels increase in phase of acute inflammation. ^[3]^ Hence, any women with signs or symptoms of acute inflammation in the last 7 days were excluded from the study.

Data were analysed using SPSS version 22. Chi-square was used to know the relation between variables. Formal approval was obtained from the District Authority and Ethical clearance was sought from the Institutional Ethics Committee at Indian Institute of Public Health Gandhinagar.

## Results

A total of 258 women were enrolled from the PHC. The overall mean age of the study participants was 25.3 years which was almost similar in the group of AN and PN women (25.7 and 25.1 years respectively). (Table 1)

**Table 1.** Age distribution of the in Antenatal (AN) and Postnatal (PN) women.

|  | **AN women (N=188)** | **PN women (N=70)** | **Overall (N=258)** |
| --- | --- | --- | --- |
| **Mean Age** | 25.7 ± 3.7 years | 25.1 ± 3.9 years | 25.3 ± 3.8 years |

Overall, Anaemia (low Hb) and ID (low s. ferritin) was observed in 65.9% and 27.1% respectively. Out of anaemic participants, about 38.2% reported ID while 61.8% had normal s. ferritin (i.e. NIDA). The difference in s. ferritin and Hb concentration among anemic and non-anaemic was statistically significant (□2=31.08, df=1, P<0.0001) (Table 2).

**Table 2.** Relation between Iron deficiency and Anemia in all patients (N=258)

| **Hb gm% status** | **S. ferritin status** |  | **Total** |
| --- | --- | --- | --- |
|  | **Deficient S. Iron** | **Normal S. Iron** |  |
| Anemia | 65 (38.2%) | 105 (61.8%) | 170 (100.0%) |
| Normal Hb | 5 (5.7%) | 83 (94.3%) | 88 (100.0%) |
| **Total** | **70 (27.1%)** | **188 (72.9%)** | **258 (100.0%)** |
| $\chi^2=31.08$, df=1, $P<0.0001$ | | | |

Among all study participants, IDA was observed in 25% while 41% had NIDA. The proportion of IDA was higher in AN woman (28%) than PN women (17%) while the proportion of NIDA was almost similar among AN and PN women (41% and 40% respectively). Overall, only 2% of them were at risk of developing anaemia as they have iron deficiency but no anaemia (Figure 1).

**Figure 1:**
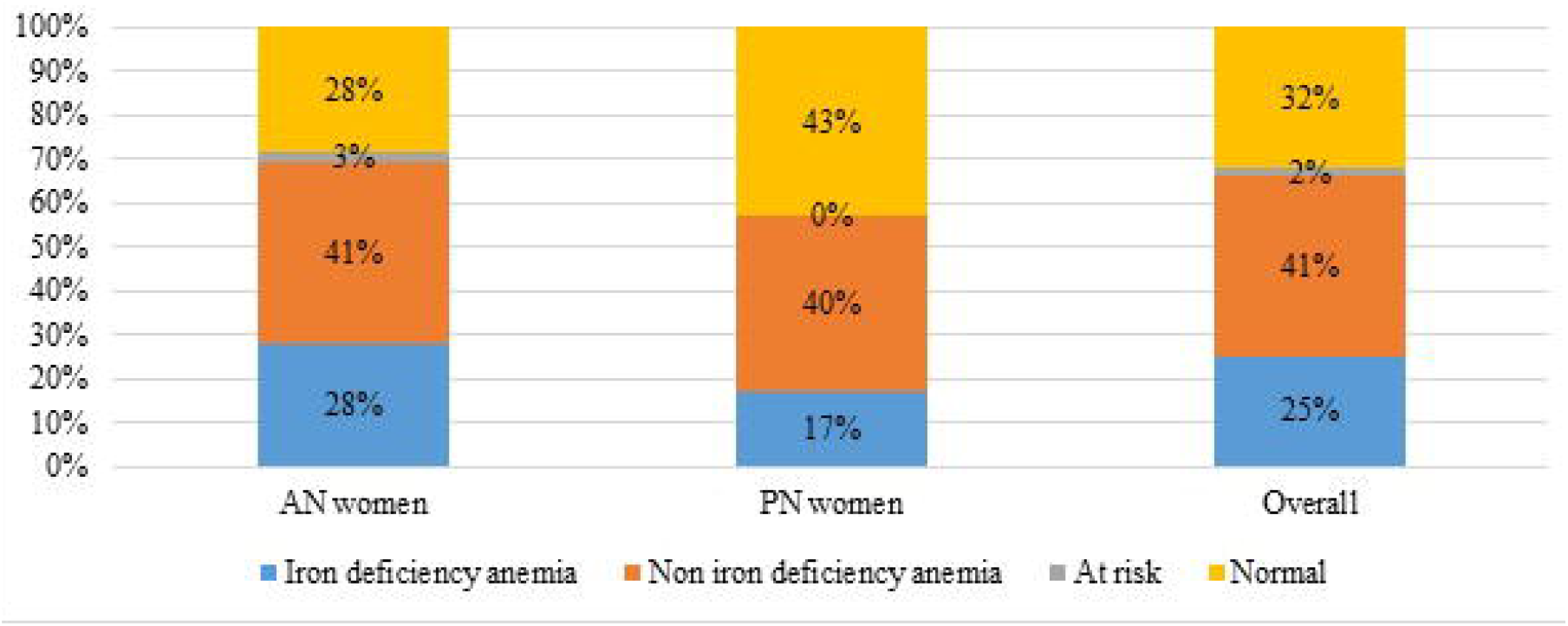
Anemia Vs Iron deficiency in participants.

Out of total participants, 73.2% were AN women and 26.8% were PN women. Anaemia was reported 69.1% in AN women and 57.1% in PN women. The ID was reported higher (30.9%) in AN women than PN women (17.1%). Amongst all anaemic participants, IDA was 40.8% in AN women and 30% in PN women whereas NIDA was observed to be 59.2% and 70% among AN and PN women respectively. (Table 3) The comparative profile of s. ferritin and Hb concentration among AN and PN women are presented in the Table 3. The difference in s. ferritin and Hb concentration among anaemic and non-anaemic group of AN women were found statistically significant (□2=19.4, df=1, P<0.0001).

**Table 3.** Relation between Iron deficiency and Anemia in AN (N=188) and PN women (N=70)

| **Hb gm% status in AN women** | **S. ferritin status** |  | **Total** |
| --- | --- | --- | --- |
|  | **Deficient S. Iron** | **Normal S. Iron** |  |
| Anemia | 53 (40.8%) | 77 (59.2%) | 170 (100.0%) |
| Normal Hb | 5 (8.6%) | 53 (91.4%) | 88 (100.0%) |
| **Total** | **58 (30.9%)** | **130 (69.1%)** | **188 (100.0%)** |
| $\chi^2=19.4$, df=1, $P<0.0001$ | | | |
| **Hb gm% status in PN women** | **S. ferritin status** |  | **Total** |
|  | **Deficient S. Iron** | **Normal S. Iron** |  |
| Anemia | 12 (30.0%) | 28 (70.0%) | 40 (100.0%) |
| Normal Hb | 0 (0.0%) | 30 (100.0%) | 30 (100.0%) |
| **Total** | **12 (17.1%)** | **58 (82.9%)** | **70 (100.0%)** |
| $\chi^2=NA$ | | | |

The prevalence of anaemia, as well as IDA decreased from the first to the third trimester; 50%, 40.7% and 39.7% in first, second and third trimester respectively. The difference between these two parameters in second trimester found statistically significant (□2=17.2, df=1, P<0.0001) while it was not significant in first trimester (□2=2.17, df=1, P=0.14). (Table 4).

**Table 4.** Trimester wise distribution of Iron deficiency and Anemia status in AN woman (N=187)

| **Hb gm% status in** | **S. ferritin status** |
| --- | --- |

| **1<sup>st</sup>-trimester women** | **Deficient S. Iron** | **Normal S. Iron** | **Total** |
| --- | --- | --- | --- |
| Anemia | 4 (50.0%) | 4 (50.0%) | 8 (100.0%) |
| Normal Hb | 2 (18.2%) | 9 (81.8%) | 11 (100.0%) |
| **Total** | **6 (31.6%)** | **13 (68.4%)** | **19 (100.0%)** |
| □ 2=2.17, df=1, P=0.14 |  |  |  |
| **Hb gm% status in** | **S. ferritin status** |  | **Total** |
| **2<sup>nd</sup>-trimester women** | **Deficient S. Iron** | **Normal S. Iron** |  |
| Anaemia | 24 (40.7%) | 35 (59.3%) | 59 (100.0%) |
| Normal Hb | 3 (21.4%) | 11 (78.6%) | 14 (100.0%) |
| **Total** | **27 (37.0%)** | **46 (63.0%)** | **73 (100.0%)** |
| □ 2=17.2, df=1, P<0.0001 |  |  |  |
| **Hb gm% status in** | **S. ferritin status** |  | **Total** |
| **3<sup>rd</sup>-trimester women** | **Deficient S. Iron** | **Normal S. Iron** |  |
| Anaemia | 25 (39.7%) | 38 (60.3%) | 63 (100.0%) |
| Normal Hb | 0 (0.0%) | 32 (100.0%) | 32 (100.0%) |
| **Total** | **25 (26.3%)** | **70 (73.7%)** | **95 (100.0%)** |
| □ 2 -- NA |  |  |  |

## Discussion

The present study was conducted to know the relation between haemoglobin and s. ferritin in pregnant and lactating women.

About 27.1% of the study participant had ID (low s. ferritin) while 38.2% of them were screened as anemia (low Hb) in the present study which is within the range of 19% to 61% reported in various studies conducted in low and middle income countries.^[11]^

The prevalence of NIDA was significantly higher than IDA in all participants as well as in AN and PN women group. In a review of European iron-status studies in women,^[12]^ ID was reported in 25–77% of pregnant women, and IDA was reported in 6–30% of pregnant women. A study conducted among AN woman in tertiary care center of India reported 34% IDA and 14% NIDA. ^[13]^ Muthukumaran et al ^[14]^ reported IDA proportion higher (60%) than the present study (40%) in the third trimester AN woman. The proportion of IDA was lower than NIDA in second trimester among anemic women. Prophylactic oral iron consumption during the second and third trimester might be the reason behind this variation.

As per National guidelines, the screening of anemia is being carried out among AN and PN women by haemoglobin estimation while no screening is being carried out for iron deficiency in India. Comprehensive National Nutrition survey 2016-18 (CNNS 2016-18), had reported higher prevalence of the NIDA (16.9%) than IDA (12%) among adolescent (age 10 – 19 years) by using s. ferritin as a screening tool and also stated ID as the strongest predictor of Anemia among adolescent.^[15]^ Similar to that large scale survey of AN, as well as PN women by s. ferritin can be useful to know the burden of the IDA as well as NIDA and focus can be prioritize for anemia control.

Various preventive measures are being carried out to address nutritional anemia includes food fortification and food diversity but for the treatment of anemia especially moderate to severe, oral and parenteral iron along with folate are indicated in second and third trimester irrespective of iron status of the body.^[4]^ Due to physiological depletion in iron storage, iron supplements are mandatory in the pregnancy period.^[16]^ As observed in the present study, a large number of AN mothers (59.2%) were anemic might be due to causes other than the ID.^[17]^ Even globally it was evidenced that reason behind half of the anemic patients was other than ID. ^[2]^ Thus, in context to nutritional anemia, other micronutrient deficiencies needed to be assessed and addressed for tackling maternal anemia effectively.

## Conclusion

Iron Deficiency Anemia is still a public health problem. However, the proportion of Non-Iron Deficiency Anemia observed in the present study is alarming that ought to be addressed. Two out of every three women are anaemic; one out of four were anaemic with depleted iron storage. Importantly, two out of five women had anaemia but iron storage was sufficient. To achieve the ambitious National nutrition target 2025 – 50% reduction of anaemia in women of reproductive age, strategy to prevent and correct anaemia must include screening for iron and non-iron deficiency anaemia and follow appropriate treatment protocol for both types of anaemia.

## Data Availability

Available on written request.

## Acknowledgements

The exercise is part of a comprehensive baseline assessment under Project Tushti, a three-year initiative (2019 – 2022) supported through Nayara Energy. The project aims to address malnutrition in Devbhumi Dwarka in collaboration with the Department of Women and Child Development through ICDS program and the Health and Family Welfare Department, Gujarat

## Funding statement

The authors declare that this study received funding from Nayara Energy. The funder had no role in study design, data collection and analysis, decision to publish, or preparation of the manuscript.

## Conflict of interest

All authors declare no conflict of interest.

## Notes

### Competing Interest Statement

The authors have declared no competing interest.

### Clinical Trial

NA

### Author Declarations

Institutional Ethics committee, Indian Institute of public Health Gandhinagar

